# Discordant Care as a Computable Phenotype for Real-Time Detection of Routine Protocol Completion Without Cognitive Patient Engagement in the ICU: Retrospective Cohort Study

**DOI:** 10.64898/2026.02.24.26347021

**Authors:** Greg Born

## Abstract

**Background:** Quality measurement in intensive care emphasizes task completion—whether assessments were documented and protocols followed. Electronic health record (EHR) systems capture these signals in real time, yet current metrics cannot distinguish task completion from cognitive clinical engagement. A prior analysis demonstrated that omission of orientation assessment predicted a 4.29-fold increase in hospital mortality among low-acuity ICU patients [1]. Whether combining this marker with routine task-completion data yields a computable phenotype with independent prognostic value has not been studied.

**Objective:** To define, validate, and characterize “discordant care”—a computable EHR phenotype defined as completion of ≥6 of 8 routine nursing assessments without orientation assessment documentation—as a predictor of hospital mortality, distinguishing patient-level confounding from care process signal.

**Methods:** Retrospective cohort study using MIMIC-IV v3.1 (2008–2022), including 46,004 adult ICU stays with SOFA scores 0-2 and length of stay ≥24 hours in non-neurological ICUs. Primary exposure: discordant care, computed from structured nursing flowsheet data within 24 hours of admission. Primary outcome: hospital mortality. Progressive covariate adjustment included mechanical ventilation, sedation, and diagnosis.

**Results:** Discordant care was present in 8891 patients (19.3%), with 69.7% mechanically ventilated versus 25.3% of concordant patients. Two overlapping signals were identified: a patient-level signal driven by ventilation/sedation (full adjustment OR 1.19, 95% CI 1.09–1.30) and a care process signal in non-ventilated patients (OR 2.14, 1.87–2.44; N=30,314). Among non-ventilated SOFA 0 patients, OR was 2.60 (2.13–3.18; N=16,295). The signal was present across all 7 major diagnosis categories. Quantitative bias analysis indicated unmeasured delirium could attenuate but likely not fully explain the non-ventilated signal.

**Conclusions:** Discordant care identifies two phenomena: a patient-level signal from ventilation/sedation and a care process signal where assessable patients receive routine care without cognitive engagement (OR 2.14–2.60). This care process signal is invisible to existing quality metrics and detectable in real time. Prospective validation with systematic delirium screening is needed.

## INTRODUCTION

### Background

Quality measurement in intensive care has long focused on task completion: were vital signs documented, were assessments performed, were protocols followed [2,3]. EHR systems generate continuous, structured records of these task-completion events, and automated quality dashboards increasingly rely on this documentation to measure care delivery [4]. This approach implicitly assumes that completing the right tasks leads to good outcomes.

However, task completion and clinical thinking are not the same thing [5], and EHR-derived metrics capturing the former may fail to capture the latter. A clinician can complete every protocol-driven assessment—repositioning, lung auscultation, skin checks, edema monitoring—without engaging the patient in dialogue or evaluating cognitive status. The distinction between assessments that can be performed on a passive body and those requiring interactive patient engagement represents a gap in current EHR-based surveillance. Diagnostic errors remain a leading cause of preventable harm in US hospitals [17], and may go undetected when quality metrics reward documentation completeness without measuring whether clinicians are cognitively engaging with patients.

### Prior Work

Prior work using the same database demonstrated that failure to document orientation assessment in the first 24 hours of ICU admission is associated with a 4.29-fold increase in mortality among low-acuity patients (SOFA 0-2), with an E-value of 8.0 [1]. A systematic screen of 14 routine ICU nursing assessments revealed that orientation is the only assessment where omission predicts increased mortality; all others show the reverse pattern [1]. This differential signal suggests a distinction between routine protocol-driven assessments (repositioning, lung auscultation, skin checks) that can be performed without patient dialogue, and assessments requiring interactive cognitive engagement (orientation) where the clinician must converse with the patient.

When EHR data shows a patient receiving extensive routine care but no interactive assessment, it may indicate care process fragmentation—or that the patient is mechanically ventilated or deeply sedated and cannot participate.

### Study Objectives

We term this EHR-detectable pattern “discordant care” and define it as a computable phenotype derived entirely from structured nursing flowsheet data. We test the hypothesis that discordant care independently predicts mortality and distinguish between patient-level confounding (mechanical ventilation, sedation) and care process signal by directly measuring ventilation status. We evaluate the phenotype’s discriminative performance as a potential real-time clinical decision support trigger.

## METHODS

### Study Design and Population

We conducted a retrospective cohort study using MIMIC-IV v3.1 [6], accessed via Google BigQuery. We included adult patients (≥18 years) with ICU length of stay of 24 hours or longer, SOFA scores 0-2 [7], admitted to non-neurological ICU units. First stay per admission was retained. The final cohort comprised 46,004 ICU stays (94,458 total → 74,829 length of stay ≥24 hours → 67,485 non-neuro → 55,294 SOFA ≤2 → 46,004 first stays).

### Computable Phenotype: Discordant Care

The discordant care phenotype is computed from 2 structured data elements available in real time from nursing flowsheets.

First, a “routine care score” (RCS) counts 8 routine nursing assessments documented within 24 hours of ICU admission: repositioning/turning, cough/secretion assessment, lung sounds, Richmond Agitation-Sedation Scale (RASS), skin assessment, Braden scale, edema assessment, and abdominal assessment. These are assessments that can be performed on a passive or unresponsive patient without interactive dialogue.

Second, cognitive engagement was operationalized as orientation assessment documentation (chartevents items 228394, 228395, 228396, 223898, 229381). This assessment requires the clinician to engage the patient in verbal interaction to evaluate orientation to person, place, and time.

*Discordant care* was defined as RCS of 6 or higher with no orientation documentation. This definition captures patients receiving extensive routine protocol-driven care without any documented interactive cognitive engagement. Sensitivity analyses confirmed stability across all RCS thresholds (Multimedia Appendix 3).

### Outcome and Covariates

The primary outcome was hospital mortality. Base covariates included age, sex, SOFA score, Charlson Comorbidity Index [8], comfort care status, night shift admission, weekend admission, and ICU unit type.

### Ventilation, Sedation, and Diagnosis Assessment

Mechanical ventilation was identified from ventilator mode documentation (chartevents itemIDs 223849, 228640). Broader ventilation indicators based on monitoring items were rejected due to 99.9% prevalence. Deep sedation was defined as any RASS of −3 or lower within 24 hours (text-parsing artifacts corrected). Confusion Assessment Method for the ICU (CAM-ICU) delirium assessment was documented in only 1037 patients (2.3%), precluding subgroup analysis. Primary diagnosis was categorized from ICD codes (sequence number 1) into cardiac, trauma/injury, sepsis/infection, respiratory, gastrointestinal, cancer, endocrine/metabolic, renal, postprocedural, and other.

### Statistical Analysis

#### Primary analysis

Multivariable logistic regression adjusted for all base covariates. *Progressive model adjustment:* 4 nested models adding ventilation, diagnosis, and deep sedation sequentially. *Non-ventilated subgroup analyses:* prespecified restrictions among patients without ventilator mode documentation, including SOFA-stratified and awake (RASS > −3) subgroups. *Inverse probability of treatment weighting (IPTW):* stabilized weights trimmed at the 1st/99th percentiles [9]. *E-values* computed per VanderWeele and Ding [10]. *Quantitative bias analysis for delirium:* bias factors computed under ranges of delirium prevalence (10%-30%), delirium-mortality OR (2.0–3.0), and probability of absent orientation documentation if delirious (70%-90%).

#### Discriminative performance

AUROC was compared between models with and without the discordant care phenotype. The phenotype-alone AUROC was compared against SOFA-alone AUROC. Clinical utility metrics including sensitivity, specificity, positive predictive value (PPV), negative predictive value (NPV), crude and adjusted number needed to screen (NNS) via marginal standardization were computed.

#### Additional analyses

Diagnosis-stratified models, dose-response by RCS, subgroup analyses with Benjamini-Hochberg false discovery rate (FDR) correction, and ventilation × discordant care interaction testing.

All analyses used Python 3.12 with statsmodels 0.14, scikit-learn 1.5, and scipy 1.14. Two-sided P<.05 was considered significant.

### Ethical Considerations

This study used the MIMIC-IV database, a deidentified publicly available critical care dataset. Access was obtained through the PhysioNet Credentialed Data Use Agreement, which requires completion of a recognized human subjects training course. Because the data are deidentified in accordance with the Health Insurance Portability and Accountability Act Safe Harbor standard, this research does not constitute human subjects research and institutional review board approval was not required. No informed consent was necessary as all data were retrospective and deidentified.

## RESULTS

### Study Population and Primary Analysis

Of 46,004 patients, 8891 (19.3%) met criteria for discordant care (Figure 3). Overall mortality was 9.3%. Discordant patients had higher unadjusted mortality (14.4% vs 8.1%).

Table 1 presents baseline characteristics. Mechanical ventilation was markedly asymmetric: 69.7% of discordant versus 25.3% of concordant patients were ventilated, indicating two overlapping phenomena captured by the phenotype.

**TABLE 1.** Baseline Characteristics by Discordant Care Status.

| Characteristic | Discordant (N=8891) | Concordant (N=37,113) | SMD |
| --- | --- | --- | --- |
| Age, years, mean (SD) | 62.3 (16.5) | 63.2 (16.9) | –0.053 |
| Male sex, n (%) | 5270 (59.3) | 20,136 (54.3) | 0.101 |
| SOFA score, mean (SD) | 0.8 (0.8) | 0.6 (0.7) | 0.203 |
| Charlson index, mean (SD) | 1.0 (1.4) | 0.7 (1.3) | 0.216 |
| Comfort care, n (%) | 416 (4.7) | 2226 (6.0) | –0.059 |
| Night shift, n (%) | 3833 (43.1) | 16,883 (45.5) | –0.048 |
| Weekend, n (%) | 2491 (28.0) | 10,552 (28.4) | –0.009 |
| **Ventilated, n (%)** | **6196 (69.7)** | **9394 (25.3)** | — |
| **Deep sedation, n (%)** | **2179 (24.5)** | **1746 (4.7)** | — |
| **ICU Unit** |  |  |  |
| MICU | 1767 (19.9) | 8767 (23.6) |  |
| SICU | 1410 (15.9) | 6214 (16.7) |  |
| CVICU | 2308 (26.0) | 4856 (13.1) |  |
| CCU | 708 (8.0) | 5444 (14.7) |  |
| TSICU | 1466 (16.5) | 4705 (12.7) |  |
| MICU/SICU | 1151 (12.9) | 7008 (18.9) |  |
| Other | 81 (0.9) | 119 (0.3) |  |
| **Hospital mortality, n (%)** | **1279 (14.4)** | **3016 (8.1)** |  |

After base adjustment, discordant care was strongly associated with mortality (OR 2.27; 95% CI 2.11–2.45; *P*<.001; Table 2). IPTW confirmed the finding (OR 2.20; 2.06–2.36; all postweighting standardized mean differences [SMDs] <0.07). E-value: 3.97 (CI lower bound 3.63).

**TABLE 2.** Multivariable Logistic Regression—Primary Model.

| Variable | Adjusted OR | 95% CI | P value |
| --- | --- | --- | --- |
| **Discordant care** | **2.27** | **2.11–2.45** | **<.001** |
| Age (per year) | 1.02 | 1.02–1.02 | <.001 |
| Male sex | 1.05 | 0.98–1.13 | .13 |
| SOFA (per unit) | 1.17 | 1.12–1.22 | <.001 |
| Charlson (per unit) | 1.17 | 1.14–1.19 | <.001 |
| Comfort care | 5.55 | 5.06–6.09 | <.001 |
| Night shift | 1.13 | 1.05–1.21 | <.001 |
| Weekend | 1.00 | 0.93–1.08 | .95 |

*Adjusted for ICU unit type (6 categories). IPTW: OR 2.20 (2.06–2.36), all SMDs <0.07 (Multimedia Appendix 2). E-value: 3.97 (lower bound 3.63).*

### Ventilation, Sedation, and Diagnosis Confounding

Ventilator mode was present in 33.9% of the cohort but markedly asymmetric: 69.7% of discordant versus 25.3% of concordant patients. Among SOFA 0, 72.3% of discordant versus 22.3% of concordant were ventilated. Deep sedation was present in 24.5% of discordant versus 4.7% of concordant patients (Table 3, Panel E).

**TABLE 3.** Progressive Adjustment and Subgroup Analyses.

**Panel A: Progressive Model Adjustment (Full Cohort)**
| Model | OR | 95% CI | P value | N |
| --- | --- | --- | --- | --- |
| 1. Base covariates | 2.27 | 2.11–2.45 | $3.68 \times 10^{-102}$ | 46,004 |
| 2. Base + ventilation | 1.63 | 1.50–1.77 | $4.20 \times 10^{-30}$ | 46,004 |
| 3. Base + diagnosis | 2.21 | 2.04–2.38 | $1.31 \times 10^{-89}$ | 46,004 |
| **4. Full (vent+dx+sed)** | **1.19** | **1.09–1.30** | **$8.71 \times 10^{-5}$** | 46,004 |

**Panel B: Non-Ventilated Subgroups**
| Subgroup | OR | 95% CI | P value | N |
| --- | --- | --- | --- | --- |
| Non-vent full cohort | 2.14 | 1.87–2.44 | $6.74 \times 10^{-29}$ | 30,314 |
| Non-vent + dx adj | 1.90 | 1.66–2.18 | $9.86 \times 10^{-21}$ | 30,314 |
| Non-vent SOFA 0 | 2.60 | 2.13–3.18 | $8.85 \times 10^{-21}$ | 16,295 |
| Non-vent SOFA 0 + act + GCS<sup>a</sup> | 2.77 | 2.25–3.40 | $7.49 \times 10^{-22}$ | 14,874 |
| Non-vent awake (RASS>–3) | 1.57 | 1.34–1.83 | $1.34 \times 10^{-8}$ | 29,598 |
| Non-vent awake SOFA 0 | 1.59 | 1.23–2.04 | $3.46 \times 10^{-4}$ | 15,997 |

Panel C: SOFA-Stratified Non-Ventilated
| SOFA | OR | 95% CI | P value | N |
| --- | --- | --- | --- | --- |
| 0 | 2.60 | 2.13–3.18 | $8.85 \times 10^{-21}$ | 16,295 |
| 1 | 1.95 | 1.53–2.49 | $8.24 \times 10^{-8}$ | 9328 |
| 2 | 1.58 | 1.21–2.06 | $7.15 \times 10^{-4}$ | 4691 |

Panel D: Ventilated (Comparison)
| Subgroup | OR | 95% CI | P value | N |
| --- | --- | --- | --- | --- |
| Ventilated full | 1.45 | 1.31–1.61 | $5.54 \times 10^{-12}$ | 15,490 |
| Ventilated SOFA 0 | 1.63 | 1.40–1.90 | $1.81 \times 10^{-10}$ | 7233 |

Panel E: Ventilation/Sedation Prevalence
| Group | N | Vent % | Deep Sed % |
| --- | --- | --- | --- |
| Full cohort | 46,004 | 33.9 | 8.5 |
| Discordant | 8891 | 69.7 | 24.5 |
| Concordant | 37,113 | 25.3 | 4.7 |
| Discordant SOFA 0 | 4066 | 72.3 | 26.6 |
| Concordant SOFA 0 | 19,604 | 22.3 | 3.9 |

<sup>a</sup>Conditioning on postexposure variables may introduce collider bias; interpret as descriptive. SOFA × discordant interaction: $P < .001$ . Ventilation × discordant interaction: $P = 7.58 \times 10^{-10}$ (implied OR 2.26 non-ventilated vs 1.34 ventilated).

Progressive adjustment revealed ventilation as the dominant confounder: adding ventilation reduced the OR from 2.27 to 1.63 (1.50–1.77). Diagnosis alone had minimal impact (OR 2.21). Full adjustment yielded OR 1.19 (1.09–1.30; *P*=8.71×10⁻⁵; Table 3, Panel A). A formal ventilation × discordant care interaction test was highly significant (*P*=7.58×10⁻¹⁰), confirming that the association differs by ventilation status: the implied OR was 2.26 in non-ventilated patients versus 1.34 in ventilated patients. CAM-ICU was too sparse for analysis (2.3% assessed; 0.10% of discordant patients).

### Non-Ventilated Subgroup Analysis

Among non-ventilated patients (N=30,314), discordant mortality was 12.7% versus 6.9%; adjusted OR 2.14 (1.87–2.44; *P*=6.74×10⁻²⁹). With diagnosis adjustment: OR 1.90 (1.66–2.18).

Among non-ventilated SOFA 0 patients (N=16,295), mortality was 13.3% versus 5.3%; OR 2.60 (2.13–3.18). Among non-ventilated SOFA 0 with activity and Glasgow Coma Scale (GCS) documentation (N=14,874), OR was 2.77 (2.25–3.40)—though this estimate may be affected by collider bias from conditioning on postexposure variables.

Among non-ventilated awake patients (RASS > −3; N=29,598), OR was 1.57 (1.34–1.83). SOFA-stratified non-ventilated: SOFA 0 (OR 2.60), SOFA 1 (OR 1.95), SOFA 2 (OR 1.58; Table 3, Panels B and C). Ventilated patients showed weaker associations (OR 1.45 full; OR 1.63 SOFA 0; Table 3, Panel D). Among discordant patients at SOFA 0, pain assessment—another interactive, non-RCS assessment—was documented in 45.9% versus 66.8% of concordant patients (difference 20.9 percentage points), suggesting a broader pattern of reduced interactive assessment beyond orientation alone.

### Diagnosis-Stratified Analysis

The signal was present across all 7 major categories: cardiac (OR 3.55; N=12,282), trauma/injury (OR 3.30), other (OR 2.80), gastrointestinal (OR 2.03), respiratory (OR 1.70), sepsis/infection (OR 1.68), and cancer (OR 1.41; Table 4). Diagnosis adjustment had minimal impact (OR 2.21 vs 2.27).

**TABLE 4.** Diagnosis-Stratified Analysis.

| Diagnosis | N | Disc Prev | OR | 95% CI | P value | Disc Mort | Conc Mort |
| --- | --- | --- | --- | --- | --- | --- | --- |
| Cardiac | 12,282 | 20.7% | 3.55 | 2.96–4.25 | $1.85 \times 10^{-42}$ | 9.7% | 5.8% |
| Trauma | 6241 | 22.3% | 3.30 | 2.65–4.10 | $7.65 \times 10^{-27}$ | 13.4% | 5.5% |
| Other | 6668 | 21.0% | 2.80 | 2.28–3.43 | $6.17 \times 10^{-23}$ | 14.4% | 6.3% |
| GI | 4285 | 12.8% | 2.03 | 1.50–2.75 | $4.19 \times 10^{-6}$ | 12.2% | 6.7% |
| Respiratory | 4010 | 19.5% | 1.70 | 1.35–2.13 | $5.90 \times 10^{-6}$ | 17.0% | 11.8% |
| Sepsis | 4150 | 24.4% | 1.68 | 1.40–2.00 | $1.18 \times 10^{-8}$ | 26.9% | 17.5% |
| Cancer | 3137 | 15.6% | 1.41 | 1.03–1.95 | .03 | 12.7% | 11.0% |

ORs adjusted for base covariates but not ventilation within category.

### Sensitivity and Robustness Analyses

Excluding comfort care: OR 2.28 (N=43,362). Excluding the cardiovascular ICU (CVICU)—the only unit with a nonsignificant result and identical mortality in both groups (2.6% vs 2.7%)—strengthened the association: OR 2.41 (N=38,840). Among non-ventilated patients excluding CVICU: OR 2.25 (1.96–2.59; N=27,037). Among non-ventilated SOFA 0 excluding comfort care and CVICU: OR 2.82 (2.27–3.51; N=14,244; discordant mortality 13.7% vs concordant 4.6%). RCS threshold stable (range 2.21–2.28; Multimedia Appendix 3). SOFA × discordant interaction: *P*<.001. Ventilation × discordant interaction: *P*=7.58×10⁻¹⁰. Full subgroup analyses in Multimedia Appendix 4.

### Discriminative Performance and Clinical Utility

At RCS 8 (N=24,660): 16.7% mortality without orientation versus 8.5% with orientation (Figure 1; Multimedia Appendix 5). Adding the discordant care phenotype improved AUROC from 0.741 to 0.757 (Δ=0.016; *P*<.001). The phenotype alone (AUROC 0.558) outperformed SOFA alone (AUROC 0.534). Clinical utility: sensitivity 29.8%, specificity 81.7%, PPV 14.4%, NPV 91.9%. Crude NNS 16; adjusted NNS 53. Non-ventilated crude risk difference 5.8 percentage points.

**FIGURE 1.**
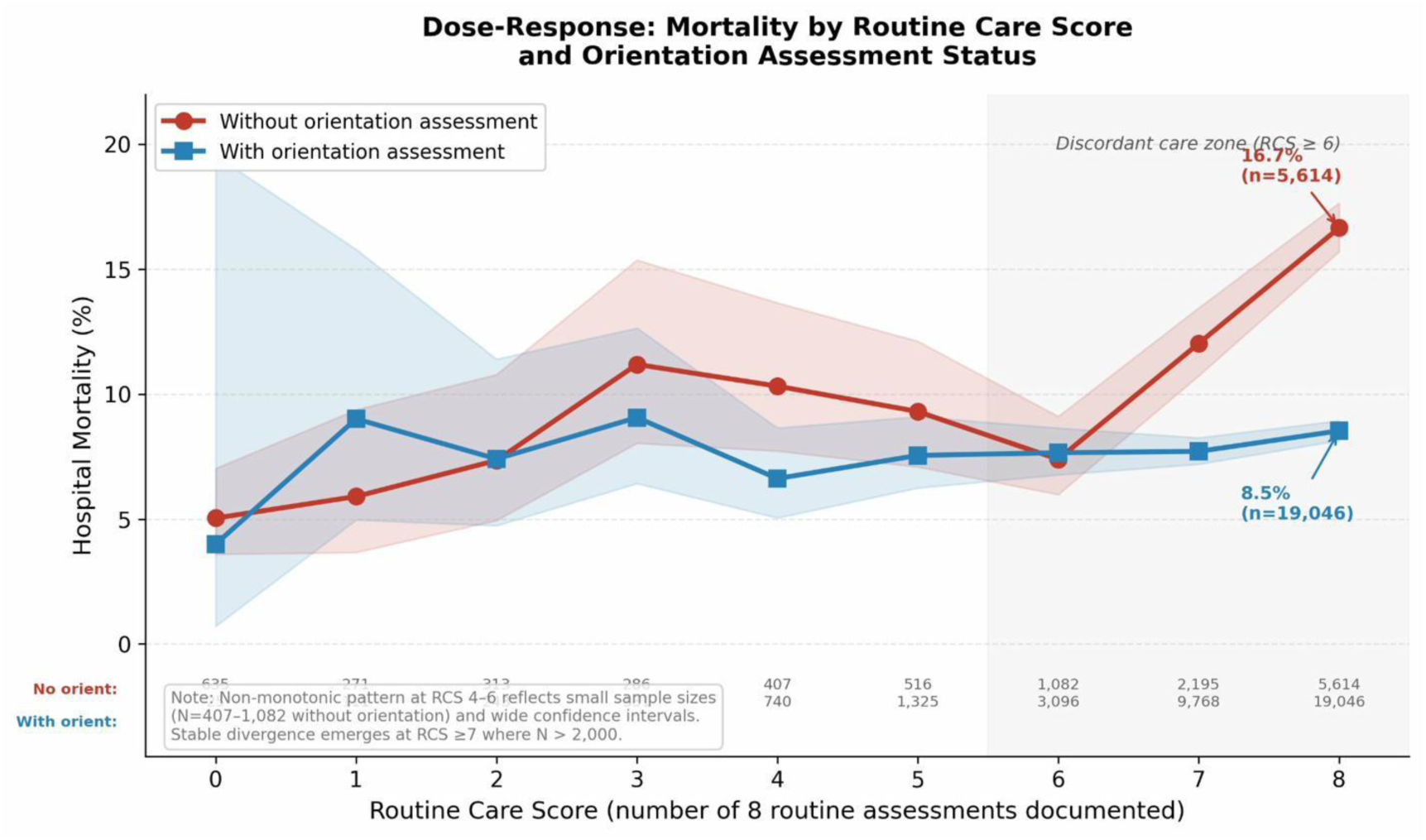
Dose-Response: Hospital Mortality by Routine Care Score and Orientation Assessment Status. Hospital mortality (%) by RCS (0-8) for patients with and without orientation documentation. Gray region marks discordant care zone (RCS ≥6). Stable divergence at RCS 7 (12.0% vs 7.7%) widens at RCS 8 (16.7% vs 8.5%, N=24,660). Non-monotonic pattern at RCS 4-6 reflects small sample sizes (N=407-1082 without orientation) and wide CIs; stable divergence emerges at RCS 7 or higher where N exceeds 2000. Raw data in Multimedia Appendix 5.

## DISCUSSION

### Principal Findings

In 46,004 low-acuity ICU patients, the discordant care phenotype was computable in real time from structured EHR data and was present in 19.3% of the cohort. Direct ventilation measurement revealed 2 overlapping phenomena captured by the phenotype: a patient-level signal (69.7% ventilated, 24.5% deeply sedated) and a care process signal in non-ventilated patients. The unadjusted OR of 2.27 attenuated to 1.19 with full adjustment. Among non-ventilated patients, OR was 2.14 (N=30,314) and 2.60 in non-ventilated SOFA 0 (N=16,295). Excluding the CVICU strengthened the association (OR 2.41). Quantitative bias analysis suggests unmeasured delirium could attenuate the non-ventilated signal substantially but likely cannot fully explain it in SOFA 0 patients.

### Two-Signal Interpretation

The first signal is patient-level: ventilated and deeply sedated patients cannot participate in orientation assessment, accounting for the majority of the crude association (OR 2.27 → 1.63 with ventilation alone). The second is a care process signal: among non-ventilated patients, discordant care identifies individuals whose care teams complete extensive assessments without interactive cognitive engagement.

The care process signal is the novel finding, though its magnitude requires careful interpretation. The non-ventilated ORs of 2.14 and 2.60 represent upper bounds, as unmeasured delirium likely accounts for a portion of the association. Nevertheless, among over 30,000 non-ventilated patients, those receiving discordant care had mortality of 12.7% versus 6.9%. The non-ventilated SOFA 0 subgroup is most defensible for the care process interpretation: no organ failure, no ventilation, and lowest expected delirium prevalence.

### Implications for EHR-Based Quality Surveillance

Traditional quality metrics ask whether the right tasks were completed [2,3]. The discordant care phenotype demonstrates that high task completion without interactive assessment is itself a risk marker—at RCS 8, patients without orientation documentation died at nearly double the rate (16.7% vs 8.5%). The missed nursing care (MISSCARE) framework [11,12] treats all omissions as failures; the data presented here show the opposite for routine assessments, whose omission reflects appropriate clinical triage. The omission that matters is the interactive assessment.

This has direct implications for EHR-based surveillance system design. Current automated quality dashboards measure documentation completeness as a proxy for care quality. The discordant care phenotype inverts this assumption: maximum documentation completeness without cognitive engagement is a risk marker, not a quality indicator. EHR alert systems could be designed to flag this pattern in real time, requiring no additional technology or documentation burden—only a new query against existing structured data.

### Mechanism and Clinical Interpretation

In non-ventilated patients, discordant care may identify unrecognized deterioration: orientation assessment requires conversational interaction that may reveal subtle mentation changes preceding decompensation. It may also reflect care process fragmentation where protocol-driven tasks proceed without higher-order interactive assessment. This interpretation is supported by the finding that pain assessment—another interactive, non-RCS assessment—was documented 20.9 percentage points less often in discordant patients at SOFA 0 (45.9% vs 66.8%).

The stronger signal at SOFA 0 compared with SOFA 2 suggests discordant care is most consequential in patients who appear well, where it may be the primary mechanism by which deterioration goes undetected.

### Alternative Explanations

#### Ventilation and sedation

Directly addressed: 69.7% of discordant patients were ventilated, and ventilation is the dominant confounder (OR: 2.27 → 1.63). A formal interaction test confirmed the discordant care effect differs significantly by ventilation status (interaction *P*=7.58×10⁻¹⁰; implied OR 2.26 non-ventilated vs 1.34 ventilated). Among non-ventilated patients excluding CVICU (N=27,037), the OR was 2.25 (1.96–2.59), eliminating both the ventilation confounder and the unit-workflow artifact simultaneously. Among non-ventilated SOFA 0 patients with documented activity and GCS (OR 2.77; N=14,874), the signal persists in a population with multiple indicators of assessability—though conditioning on these postexposure variables may introduce collider bias, and the progressive strengthening (2.14 → 2.60 → 2.77) should be interpreted as descriptive characterization rather than causal evidence against confounding.

#### Unit-specific workflows

The CVICU had the highest discordant prevalence (32.2%) and identical mortality in both groups (2.6% vs 2.7%). Excluding CVICU strengthened the association (OR 2.41), and the signal was significant in all 5 other major ICU types (ORs 1.64–3.26). The CVICU finding likely reflects postoperative cardiac surgery workflows where routine assessments are protocolized and orientation assessment is less clinically relevant.

#### Diagnosis and comfort care

Diagnosis adjustment had minimal impact (OR 2.21 vs 2.27). Comfort care was less prevalent in discordant patients (4.7% vs 6.0%); excluding comfort care: OR 2.28. SOFA 0 excluding comfort care and CVICU: OR 2.88 (N=20,111).

#### Delirium—the strongest remaining unmeasured confounder

CAM-ICU was assessed in only 2.3% of the cohort. Delirium is the most plausible remaining confounder: delirious patients cannot reliably undergo orientation assessment, and delirium is independently associated with ICU mortality (OR ∼2-3) [13,14].

Published literature provides empirical anchors for the delirium assumptions used in our quantitative bias analysis. The pooled prevalence of ICU delirium across 48 studies is 31% (95% CI 24%-41%) [21], with substantially higher rates in mechanically ventilated patients (up to 80%) [22]. In a prospective study of 261 non-ventilated medical ICU patients with systematic twice-daily CAM-ICU screening, delirium occurred in 48% [23]—though these patients had median APACHE II scores of 13, reflecting moderate-to-high acuity. Severity of illness is the strongest independent predictor of delirium, with each unit increase in APACHE II conferring an OR of 1.28 [24]. Our non-ventilated SOFA 0 subgroup—by definition lacking organ failure and mechanical ventilation, which are the 2 strongest delirium risk factors—would be expected to have substantially lower delirium prevalence than these reference populations. A reasonable estimate for non-ventilated SOFA 0 patients is 10%-15%, though this has not been directly measured.

Quantitative bias analysis: the E-value for the non-ventilated OR of 2.14 is 3.70. Under conservative assumptions for non-ventilated patients (15% delirium prevalence, mortality OR 2.0, 70% probability of absent orientation if delirious), the bias factor is 1.63 and the delirium-adjusted OR is approximately 1.32. Under moderate assumptions (20% prevalence, OR 2.5, 80% probability), the adjusted OR is approximately 1.05. For non-ventilated SOFA 0 (OR 2.60), where the empirical literature supports a delirium prevalence of 10%-15%, moderate assumptions yield a delirium-adjusted OR of approximately 1.30—reduced but persistent. The non-ventilated awake subgroup (OR 1.57) is most vulnerable and could be fully explained by delirium under moderate assumptions.

We interpret these bounds honestly: delirium likely accounts for a substantial portion of the non-ventilated association, and the true care process signal is smaller than the unadjusted non-ventilated ORs suggest. Complete elimination requires aggressive assumptions about delirium prevalence that exceed published estimates for comparable low-acuity populations, and the SOFA 0 subgroup provides the most buffer. Prospective studies with systematic delirium screening are essential.

### System Design Implications

In ventilated patients, the phenotype recapitulates ventilation status and has limited incremental value for clinical decision support. In non-ventilated patients, it identifies awake, breathing patients receiving extensive routine care without interactive cognitive engagement—a pattern invisible to standard quality dashboards.

#### Proposed alert workflow (Figure 2)

The discordant care phenotype triggers a structured response pathway for non-ventilated, non-CVICU ICU patients: (1) a real-time query runs continuously against nursing flowsheet data, evaluating the 8 RCS assessment fields and 5 orientation item IDs; (2) when discordant care is detected (RCS ≥6, no orientation), an alert fires to the assigned bedside nurse; (3) the nurse performs a structured cognitive status check within 2 hours—a brief, standardized assessment of orientation to person, place, and time, plus screening for acute symptom changes (pain, dyspnea, anxiety); (4) if the patient is oriented and stable, the assessment is documented and the flag is resolved; (5) if the patient is disoriented, confused, or shows new symptoms, a structured escalation occurs—CAM-ICU screening, provider notification, and reassessment of the care plan. This decision tree converts the phenotype from a passive risk marker into an actionable workflow prompt and simultaneously addresses the delirium confounder by generating systematic screening data. The intervention requires no new technology, no additional monitoring devices, and no changes to existing documentation systems—only a new query and alert rule within the existing EHR infrastructure.

**FIGURE 2.**
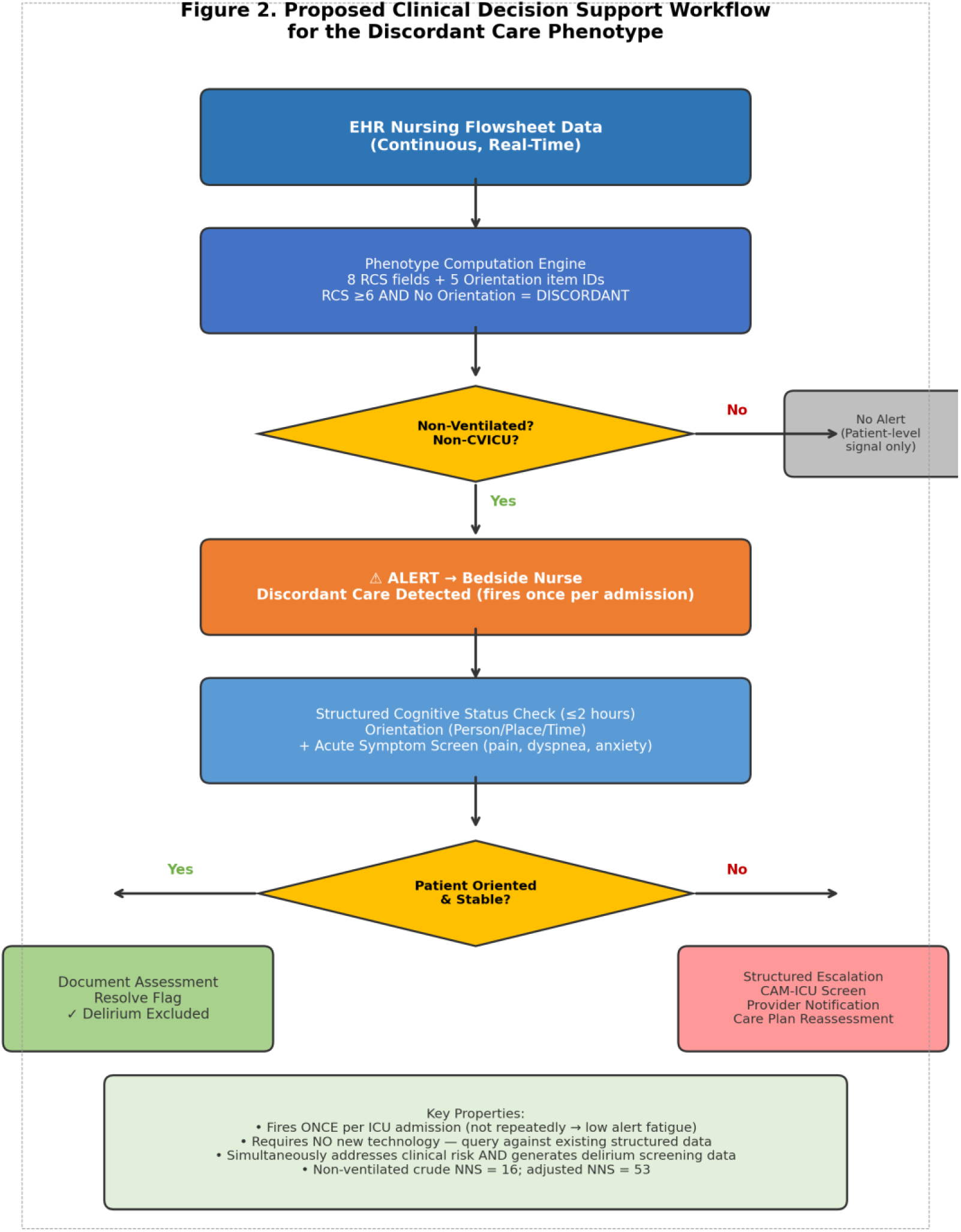
Proposed Clinical Decision Support Workflow for the Discordant Care Phenotype. Flowchart depicting the proposed real-time alert pathway. EHR nursing flowsheet data feeds continuously into the phenotype computation engine, which evaluates 8 routine care score fields and 5 orientation assessment item IDs. When the discordant pattern is detected (RCS ≥6, no orientation) in a non-ventilated, non-CVICU patient, an alert is generated to the assigned bedside nurse. The nurse performs a structured cognitive status check (orientation to person/place/time plus acute symptom screening) within 2 hours. If the patient is oriented and stable, the assessment is documented and the flag resolves. If the patient is disoriented or symptomatic, a structured escalation occurs: CAM-ICU delirium screening, provider notification, and care plan reassessment. This workflow simultaneously addresses the clinical risk and generates systematic delirium screening data to resolve the primary unmeasured confounder.

**FIGURE 3.**
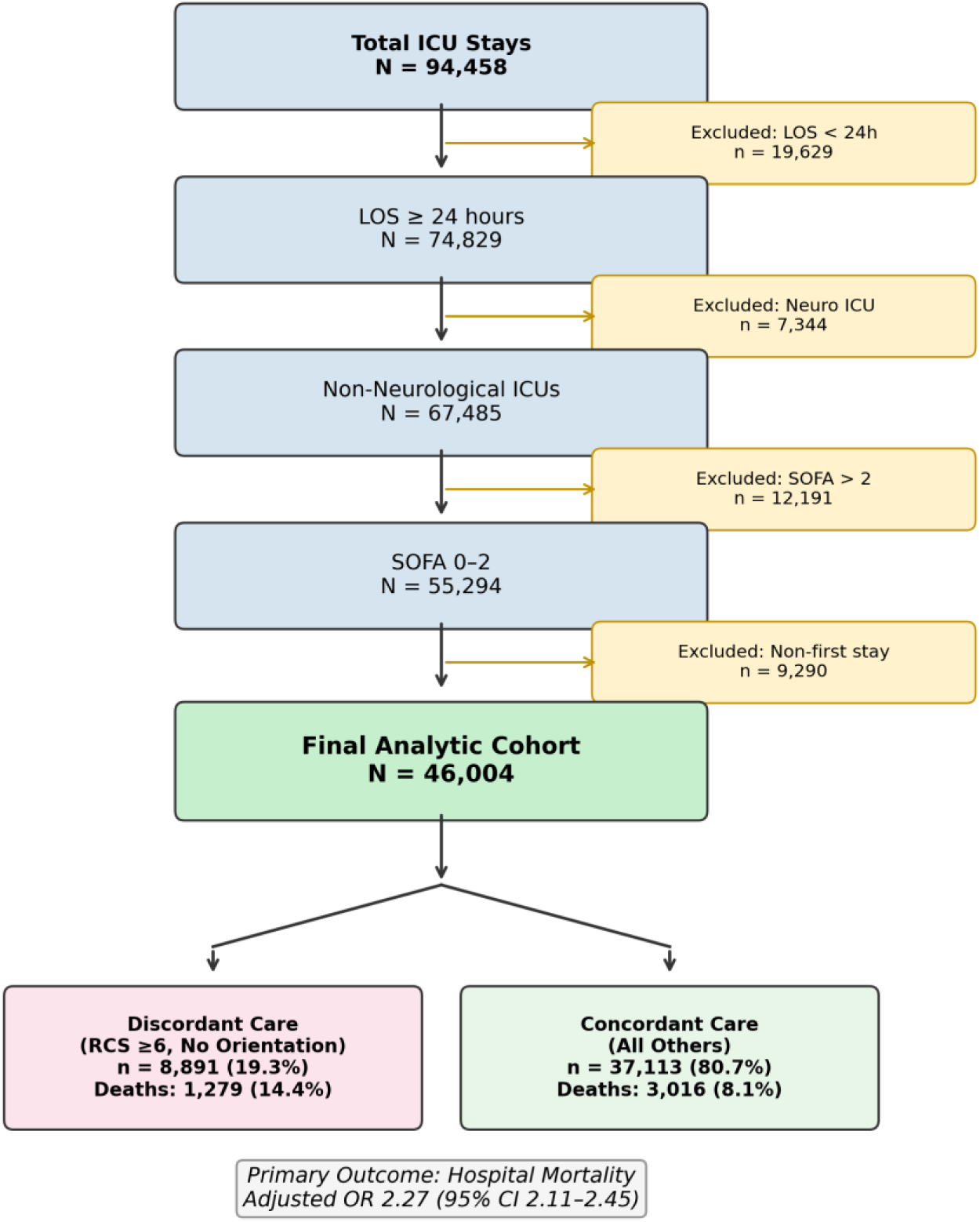
Study Flow Diagram. Flowchart depicting participant selection from 94,458 total ICU stays to the final analytic cohort of 46,004 patients, with classification into discordant (n=8,891) and concordant (n=37,113) groups. Sequential exclusions: length of stay <24 hours, neurological ICU, SOFA >2, and non-first stays.

#### Alert fatigue considerations

The phenotype has a PPV of 14.4%, meaning approximately 6 of 7 flagged patients will survive regardless. However, the flag fires once per ICU admission (not repeatedly), applies only to the 19.3% of patients meeting criteria, and triggers a low-burden intervention (brief bedside check, not a diagnostic workup). In non-ventilated patients, the crude risk difference of 5.8 percentage points and NNS of 16 compare favorably with other widely deployed clinical alerts. The adjusted NNS of 53 reflects the delirium-attenuated estimate and represents the more conservative deployment justification.

#### EHR portability

The phenotype relies on 2 categories of structured nursing flowsheet data: routine physical assessments and orientation documentation. Both are standard components of ICU nursing workflows across EHR platforms. In Epic (used by approximately 38% of US hospitals), nursing flowsheets contain structured assessment rows for neurological, respiratory, skin, and activity assessments, including orientation fields. Oracle Health (Cerner) uses a comparable flowsheet architecture with discrete assessment fields. The specific item IDs differ across platforms and institutions, but the underlying data model—structured, timestamped, binary documentation of whether a nursing assessment was performed—is universal to modern EHR systems. The primary portability challenge is not data model compatibility but institutional variation in which assessments are configured as discrete versus free-text fields. Our prior analysis of 166 eICU hospitals found that only 5% document orientation routinely [25], suggesting that the phenotype’s value may be highest in institutions with structured orientation documentation and that standardization of orientation assessment documentation is itself a quality improvement target. Multisite validation using databases with diverse EHR implementations (eICU Collaborative Research Database, 208 hospitals) is the necessary next step.

## Limitations

This study has several important limitations. First, it is a single-center retrospective study using data from one academic medical center; external validation across multiple institutions is needed. Second, we cannot determine whether orientation was performed but not documented; prospective validation with direct observation is required. Third, no causal inference is possible—discordant care may be a marker rather than a cause of elevated mortality. Fourth, MIMIC-IV date-shifting limits temporal trend analysis. Fifth, the RCS weights all 8 assessments equally, and future implementations may benefit from weighted scoring. Sixth, delirium is the strongest remaining confounder; under moderate assumptions it could attenuate the non-ventilated OR from 2.14 to approximately 1.05–1.32, and could fully explain the awake subgroup (OR 1.57); the SOFA 0 subgroup (OR 2.60) is most robust. Seventh, progressive restriction analyses condition on postexposure variables, potentially introducing collider bias; we frame these as descriptive rather than causal. Eighth, the ventilation proxy (ventilator mode) may include CPAP/BiPAP, potentially overcounting invasive ventilation.

## Conclusions

Discordant care is a computable EHR phenotype that captures a patient-level signal driven by ventilation and sedation alongside a care process signal in non-ventilated patients (OR 2.14–2.60). The care process signal is the novel contribution: assessable patients receiving routine protocol-driven care without interactive cognitive engagement have elevated mortality not fully explained by measured or plausible unmeasured confounders. The phenotype is computable in real time from existing structured nursing data, is consistent across diagnosis categories, is not driven by CVICU workflows, and outperforms SOFA alone as a discriminative marker. Prospective validation with systematic delirium screening and multisite replication are the critical next steps toward clinical deployment.

## Data Availability

This study used MIMIC-IV v3.1, publicly available at https://mimic.mit.edu/ upon completion of required training and a data use agreement. The study cohort (N=46,004) and derivation SQL are described in the Methods section. All analytical code is available upon reasonable request to the corresponding author.

https://physionet.org/content/mimiciv/3.1/

## Acknowledgments

None.

## Authors’ Contributions

GB conceived the study, designed the analysis, wrote the BigQuery SQL, performed all statistical analyses, created all figures, and wrote the manuscript.

## AI Disclosure

AI tools (Claude, Anthropic) assisted with statistical code review, literature organization, and manuscript formatting. All scientific content, study design, analysis, interpretation, and conclusions represent the author’s independent work. The author takes full responsibility for the accuracy of all reported results.

## Abbreviations

AUROC: area under the receiver operating characteristic curve
CAM-ICU: Confusion Assessment Method for the Intensive Care Unit
CVICU: cardiovascular intensive care unit
EHR: electronic health record
FDR: false discovery rate
GCS: Glasgow Coma Scale
IPTW: inverse probability of treatment weighting
MICU: medical intensive care unit
MISSCARE: missed nursing care
NNS: number needed to screen
NPV: negative predictive value
OR: odds ratio
PPV: positive predictive value
RASS: Richmond Agitation-Sedation Scale
RCS: routine care score
SICU: surgical intensive care unit
SMD: standardized mean difference
SOFA: Sequential Organ Failure Assessment
TSICU: trauma surgical intensive care unit

## Multimedia Appendices

**Multimedia Appendix 1.** STROBE checklist for cohort studies [16].

**Multimedia Appendix 2.** Post-IPTW covariate balance (Supplementary Table S2).

**Multimedia Appendix 3.** Sensitivity to routine care score threshold (Supplementary Table S3).

**Multimedia Appendix 4.** Full subgroup analyses with FDR correction (Supplementary Table S4).

**Multimedia Appendix 5.** Dose-response raw data (Supplementary Table S5).

**Multimedia Appendix 6. BigQuery SQL cohort definition and analysis code.**

**Multimedia Appendices**

*Discordant Care as a Computable Phenotype for Real-Time Detection of Routine Protocol Completion Without Cognitive Patient Engagement in the ICU: Retrospective Cohort Study*

Greg Born, MBA

## Multimedia Appendix 1. STROBE Statement Checklist for Cohort Studies

Items from the Strengthening the Reporting of Observational Studies in Epidemiology (STROBE) checklist mapped to manuscript sections.

| **Item** | **Topic** | **STROBE Recommendation** | **Location in Manuscript** |
| --- | --- | --- | --- |
| 1a | Title and abstract | Indicate the study's design with a commonly used term in the title or the abstract | Title, Abstract |
| 1b |  | Provide in the abstract an informative and balanced summary of what was done and what was found | Abstract |
| 2 | Background/rationale | Explain the scientific background and rationale for the investigation being reported | Introduction ¶1–2 |
| 3 | Objectives | State specific objectives, including any prespecified hypotheses | Introduction ¶3 |
| 4 | Study design | Present key elements of study design early in the paper | Methods: Study Design |
| 5 | Setting | Describe the setting, locations, and relevant dates, including periods of recruitment, exposure, follow-up, and data collection | Methods: Study Design |
| 6a | Participants | Give the eligibility criteria, and the sources and methods of selection of participants | Methods: Study Population |
| 6b |  | For matched studies, give matching criteria and number of exposed and unexposed | N/A (not a matched study) |
| 7 | Variables | Clearly define all outcomes, exposures, predictors, potential confounders, and effect modifiers | Methods: Exposure, Outcome, Covariates |
| 8 | Data sources/measurement | For each variable of interest, give sources of data and details of methods of assessment | Methods: Exposure (¶2–3) |
| 9 | Bias | Describe any efforts to address potential sources of bias | Methods: Statistical Analysis;<br>Discussion: Alternative Explanations |
| 10 | Study size | Explain how the study size was arrived at | Methods: Study Population; Study Flow |
| 11 | Quantitative variables | Explain how quantitative variables were handled in the analyses | Methods: Statistical Analysis |
| 12a | Statistical methods | Describe all statistical methods, including those used to control for confounding | Methods: Statistical Analysis |
| 12b |  | Describe any methods used to examine subgroups and interactions | Methods: Subgroup analyses |
| 12c |  | Explain how missing data were addressed | Methods: Covariates (SOFA, Charlson defaulted to 0 if missing) |
| 12d |  | If applicable, describe analytical methods taking account of sampling strategy | N/A |
| 12e |  | Describe any sensitivity analyses | Methods: IPTW, Threshold Sensitivity, E-values |
| 13a | Participants | Report numbers of individuals at each stage of study | Results: Study Population; Study Flow |
| 13b |  | Give reasons for non-participation at each stage | Study Flow (exclusion criteria listed) |
| 13c |  | Figure 3 (Study Flow Diagram) | Figure 3 (Study Flow Diagram) |
| 14a | Descriptive data | Give characteristics of study participants and information on exposures and potential confounders | Table 1 |
| 14b |  | Indicate number of participants with missing data for each variable of interest | Methods: Study Population (complete case analysis) |
| 15 | Outcome data | Report numbers of outcome events or summary measures | Table 1; Results ¶1 |
| 16a | Main results | Give unadjusted estimates and, if applicable, confounder-adjusted estimates and their precision | Table 2; Results: Primary Analysis |
| 16b |  | Report category boundaries when | Methods: Exposure |

| Item | Topic | STROBE Recommendation | Location in Manuscript |
| --- | --- | --- | --- |
| | | continuous variables were categorized | (RCS $\geq 6$ threshold) |
| 16c |  | If relevant, consider translating estimates of relative risk into absolute risk for a meaningful time period | Results: Clinical Utility (NNS=16) |
| 17 | Other analyses | Report other analyses done—e.g., analyses of subgroups and interactions, and sensitivity analyses | Tables 3–4; Multimedia Appendices 2–5 |
| 18 | Key results | Summarize key results with reference to study objectives | Discussion: Principal Findings |
| 19 | Limitations | Discuss limitations of the study, taking into account sources of potential bias or imprecision | Discussion: Limitations |
| 20 | Interpretation | Give a cautious overall interpretation of results considering objectives, limitations, multiplicity of analyses, results from similar studies, and other relevant evidence | Discussion: Mechanism and Interpretation |
| 21 | Generalizability | Discuss the generalizability (external validity) of the study results | Discussion: ICU as Proof of Concept |
| 22 | Funding | Give the source of funding and the role of the funders for the present study and, if applicable, for the original study on which the present article is based | Back matter: No external funding |

## Multimedia Appendix 2. Post-IPTW Covariate Balance (Table S2)

Standardized mean differences (SMD) before and after inverse probability of treatment weighting for the discordant care exposure. Propensity scores were estimated using logistic regression with all covariates. Stabilized weights were trimmed at the 1st and 99th percentiles. Target: all post-weighting SMDs <0.10.

| Covariate | Pre-weighting SMD | Post-weighting SMD | Balance Achieved |
| --- | --- | --- | --- |
| Age (years) | -0.053 | -0.006 | Yes |

| Covariate | Pre-weighting<br>SMD | Post-weighting<br>SMD | Balance<br>Achieved |
| --- | --- | --- | --- |
| Male sex | +0.101 | +0.002 | Yes |
| SOFA score | +0.203 | -0.003 | Yes |
| Charlson comorbidity<br>index | +0.216 | +0.062 | Yes |
| Comfort care status | -0.059 | -0.013 | Yes |
| Night shift admission | -0.048 | +0.022 | Yes |
| Weekend admission | -0.009 | +0.005 | Yes |

*IPTW-adjusted OR for discordant care: 2.20 (95% CI 2.06–2.36). All post-weighting SMDs <0.07, indicating excellent covariate balance across all measured confounders.*

## Multimedia Appendix 3. RCS Threshold Sensitivity Analysis (Table S3)

Adjusted odds ratios for the discordant care–mortality association across all routine care score (RCS) thresholds from ≥1 to ≥8. Discordant care is redefined at each threshold as RCS ≥ threshold with no orientation documentation. All models adjusted for age, sex, SOFA, Charlson index, comfort care, admission shift, weekend, and ICU unit type.

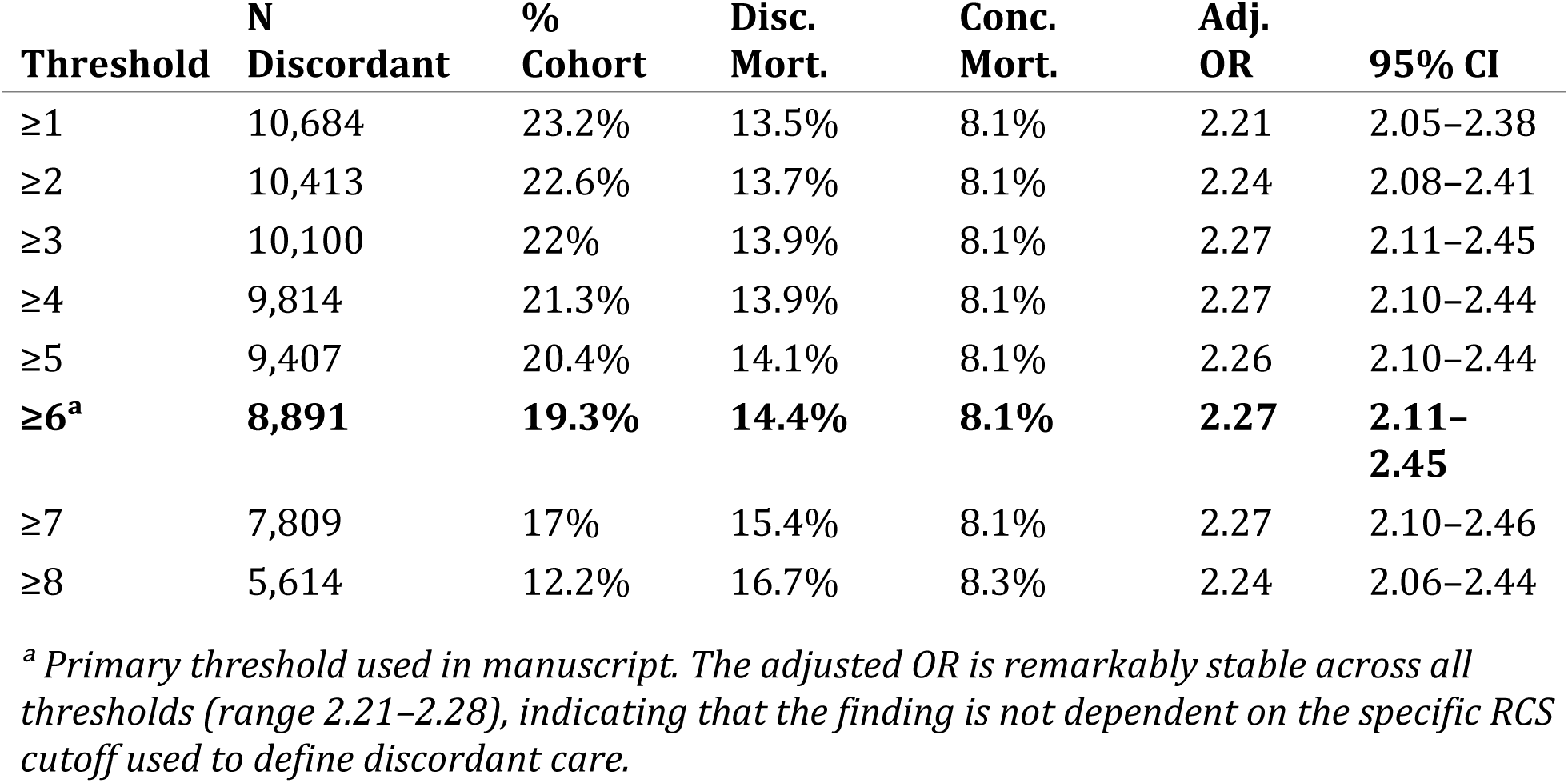

| Threshold | N<br>Discordant | %<br>Cohort | Disc.<br>Mort. | Conc.<br>Mort. | Adj.<br>OR | 95% CI |
| --- | --- | --- | --- | --- | --- | --- |
| $\geq 1$ | 10,684 | 23.2% | 13.5% | 8.1% | 2.21 | 2.05–2.38 |
| $\geq 2$ | 10,413 | 22.6% | 13.7% | 8.1% | 2.24 | 2.08–2.41 |
| $\geq 3$ | 10,100 | 22% | 13.9% | 8.1% | 2.27 | 2.11–2.45 |
| $\geq 4$ | 9,814 | 21.3% | 13.9% | 8.1% | 2.27 | 2.10–2.44 |
| $\geq 5$ | 9,407 | 20.4% | 14.1% | 8.1% | 2.26 | 2.10–2.44 |
| **$\geq 6^a$** | **8,891** | **19.3%** | **14.4%** | **8.1%** | **2.27** | **2.11–**<br>**2.45** |
| $\geq 7$ | 7,809 | 17% | 15.4% | 8.1% | 2.27 | 2.10–2.46 |
| $\geq 8$ | 5,614 | 12.2% | 16.7% | 8.3% | 2.24 | 2.06–2.44 |

<sup>a</sup> Primary threshold used in manuscript. The adjusted OR is remarkably stable across all thresholds (range 2.21–2.28), indicating that the finding is not dependent on the specific RCS cutoff used to define discordant care.

## Multimedia Appendix 4. Full Subgroup Analyses With FDR Correction (Table S4)

Adjusted odds ratios for the discordant care–mortality association across all pre-specified subgroups. Models adjusted for available covariates within each subgroup. Benjamini-Hochberg false discovery rate (FDR) correction applied across all tests. Interaction P values test effect modification by each stratifying variable in the full cohort model.

| **Subgroup** | **N** | **Disc.**<br>**Mort.** | **Conc.**<br>**Mort.** | **Adj.**<br>**OR** | **95%**<br>**CI** | **p** | **FDR** |
| --- | --- | --- | --- | --- | --- | --- | --- |
| ***SOFA Score Strata*** |  |  |  |  |  |  |  |
| SOFA 0 | 23,670 | 15.8% | 6.8% | **2.78** | 2.50–3.10 | <0.001 | ✓ |
| SOFA 1 | 14,233 | 14% | 8.9% | **2.21** | 1.92–2.54 | <0.001 | ✓ |
| SOFA 2 | 8,101 | 12.2% | 10.9% | **1.58** | 1.34–1.87 | <0.001 | ✓ |
| ***ICU Unit Type*** |  |  |  |  |  |  |  |
| Cardiac Vascular Intensive Care Unit (CVICU) | 7,164 | 2.6% | 2.7% | **1.31** | 0.93–1.83 | 0.124 | ✗ |
| Coronary Care Unit (CCU) | 6,152 | 21.3% | 7.9% | **2.97** | 2.39–3.69 | <0.001 | ✓ |
| Medical Intensive Care Unit (MICU) | 10,534 | 20.7% | 10.6% | **2.25** | 1.95–2.59 | <0.001 | ✓ |
| Medical/Surgical Intensive Care Unit (MICU/SICU) | 8,159 | 18.4% | 10.9% | **1.64** | 1.38–1.96 | <0.001 | ✓ |
| Surgical Intensive Care Unit (SICU) | 7,624 | 18.7% | 7% | **3.26** | 2.73–3.90 | <0.001 | ✓ |
| Trauma SICU (TSICU) | 6,171 | 14.1% | 6.4% | **2.92** | 2.37–3.61 | <0.001 | ✓ |
| ***Temporal Factors*** |  |  |  |  |  |  |  |
| Day shift | 25,288 | 11.8% | 7.2% | **2.46** | 2.20–2.75 | <0.001 | ✓ |
| Night shift | 20,716 | 17.7% | 9.3% | **2.21** | 1.99–2.46 | <0.001 | ✓ |
| Weekday | 32,961 | 14.1% | 8.2% | **2.23** | 2.04–2.44 | <0.001 | ✓ |
| Weekend | 13,043 | 15.1% | 8% | **2.39** | 2.04–2.71 | <0.001 | ✓ |
| ***Demographics*** |  |  |  |  |  |  |  |
| Age <65 | 22,872 | 10.8% | 5.4% | **2.38** | 2.11–2.70 | <0.001 | ✓ |
| Age ≥65 | 23,132 | 18.1% | 10.8% | **2.17** | 1.97–2.40 | <0.001 | ✓ |
| Male | 25,406 | 13.4% | 7.8% | **2.27** | 2.04–2.52 | <0.001 | ✓ |

| Subgroup | N | Disc. Mort. | Conc. Mort. | Adj. OR | 95% CI | p | FDR |
| --- | --- | --- | --- | --- | --- | --- | --- |
| Female | 20,598 | 15.9% | 8.5% | **2.28** | 2.04–2.55 | <0.001 | ✓ |
| **Comorbidity** |  |  |  |  |  |  |  |
| Charlson 0 | 30,778 | 12.1% | 7.2% | **2.57** | 2.31–2.86 | <0.001 | ✓ |
| Charlson ≥2 | 10,442 | 19.7% | 12.1% | **1.90** | 1.68–2.15 | <0.001 | ✓ |
| **Sensitivity Analysis** |  |  |  |  |  |  |  |
| Excl. comfort care | 43,362 | 12.3% | 6.5% | **2.28** | 2.10–2.48 | <0.001 | ✓ |

## Multimedia Appendix 5. Dose-Response Raw Data (Table S5)

Hospital mortality by routine care score (0–8) stratified by orientation assessment status. 95% Wilson score confidence intervals provided. Mean SOFA and age shown to characterize acuity differences between groups at each RCS level.

Panel A: Patients Without Orientation Documentation
| RCS | N | Deaths | Mortality % | 95% CI | SOFA | Age |
| --- | --- | --- | --- | --- | --- | --- |
| 0 | 635 | 32 | 5.0% | 3.6–7% | 1.04 | 62.5 |
| 1 | 271 | 16 | 5.9% | 3.7–9.4% | 0.91 | 64.0 |
| 2 | 313 | 23 | 7.3% | 4.9–10.8% | 0.81 | 63.9 |
| 3 | 286 | 32 | 11.2% | 8–15.4% | 0.85 | 63.5 |
| 4 | 407 | 42 | 10.3% | 7.7–13.7% | 0.85 | 64.3 |
| 5 | 516 | 48 | 9.3% | 7.1–12.1% | 0.93 | 62.6 |
| **6<sup>a</sup>** | **1,082** | **80** | **7.4%** | 6–9.1% | 0.96 | 64.7 |
| **7<sup>a</sup>** | **2,195** | **264** | **12.0%** | 10.7–13.5% | 0.90 | 63.4 |
| **8<sup>a</sup>** | **5,614** | **935** | **16.7%** | 15.7–17.7% | 0.71 | 61.4 |

Panel B: Patients With Orientation Documentation
| RCS | N | Deaths | Mortality % | 95% CI | SOFA | Age |
| --- | --- | --- | --- | --- | --- | --- |
| 0 | 25 | 1 | 4.0% | 0.7–19.5% | 0.68 | 61.6 |
| 1 | 111 | 10 | 9.0% | 5–15.8% | 0.53 | 62.3 |
| 2 | 243 | 18 | 7.4% | 4.7–11.4% | 0.70 | 64.5 |
| 3 | 331 | 30 | 9.1% | 6.4–12.6% | 0.69 | 64.4 |
| 4 | 740 | 49 | 6.6% | 5–8.6% | 0.66 | 62.0 |
| 5 | 1,325 | 100 | 7.5% | 6.2–9.1% | 0.68 | 64.1 |
| 6 | 3,096 | 237 | 7.7% | 6.8–8.6% | 0.68 | 63.7 |
| 7 | 9,768 | 753 | 7.7% | 7.2–8.3% | 0.62 | 63.5 |
| 8 | 19,046 | 1625 | 8.5% | 8.1–8.9% | 0.59 | 62.8 |

<sup>a</sup> Denotes RCS ≥6, qualifying as discordant care when orientation is absent. The divergence is most pronounced at RCS 7–8 where patient volumes are largest (N>2,000 without orientation). Non-monotonic dip at RCS 5–6 in the no-orientation group (9.3% and 7.4%) reflects small sample sizes and should be interpreted with caution.

## Multimedia Appendix 6. BigQuery SQL Cohort Derivation Code

Complete SQL query used to derive the N=46,004 study cohort from MIMIC-IV v3.1 via Google BigQuery. This query produces the patient-level dataset used for all analyses in this manuscript.

-- ORIGINAL QUERY THAT PRODUCED N=46,004 COHORT

-- Reconstructed from conversation history

WITH base_cohort AS (

SELECT

ie.stay_id,

ie.subject_id,

ie.hadm_id,

ie.intime AS icu_intime,

ie.outtime AS icu_outtime,

ie.first_careunit,

ie.los AS icu_los,

p.anchor_age AS age,

p.gender,

a.hospital_expire_flag,

a.admittime,

a.dischtime,

a.admission_type,

a.discharge_location,

CASE

WHEN EXTRACT(HOUR FROM ie.intime) BETWEEN 7 AND 18

THEN ‘Day’

ELSE ‘Night’

END AS admission_shift,

CASE

WHEN EXTRACT(DAYOFWEEK FROM ie.intime) IN (1, 7)

THEN ‘Weekend’

ELSE ‘Weekday’

END AS weekend_flag,

EXTRACT(YEAR FROM ie.intime) AS admission_year

FROM physionet-data.mimiciv_3_1_icu.icustays ie

INNER JOIN physionet-data.mimiciv_3_1_hosp.patients p ON ie.subject_id = p.subject_id

INNER JOIN physionet-data.mimiciv_3_1_hosp.admissions a ON ie.hadm_id = a.hadm_id

WHERE ie.los >= 1

AND p.anchor_age >= 18

),

-- Charlson comorbidity index

charlson AS (

SELECT

hadm_id,

MAX(CASE WHEN icd_code LIKE ‘I21%’ OR icd_code LIKE ‘I22%’ OR icd_code LIKE ‘410%’ OR icd_code LIKE ‘412%’ THEN 1 ELSE 0 END) AS mi,

MAX(CASE WHEN icd_code LIKE ‘I50%’ OR icd_code LIKE ‘428%’ THEN 1 ELSE 0 END) AS chf,

MAX(CASE WHEN icd_code LIKE ‘I70%’ OR icd_code LIKE ‘I71%’ OR icd_code LIKE ‘440%’ OR icd_code LIKE ‘441%’ THEN 1 ELSE 0 END) AS pvd,

MAX(CASE WHEN icd_code LIKE ‘I6%’ OR icd_code BETWEEN ‘430’ AND ‘438’ THEN 1 ELSE 0 END) AS cvd,

MAX(CASE WHEN icd_code LIKE ‘F0%’ OR icd_code LIKE ‘290%’ THEN 1 ELSE 0 END) AS dementia,

MAX(CASE WHEN icd_code LIKE ‘J4%’ OR icd_code BETWEEN ‘490’ AND ‘496’ THEN 1 ELSE 0 END) AS copd,

MAX(CASE WHEN icd_code LIKE ‘M05%’ OR icd_code LIKE ‘M06%’ OR icd_code LIKE ‘714%’ THEN 1 ELSE 0 END) AS rheum,

MAX(CASE WHEN icd_code LIKE ‘K25%’ OR icd_code LIKE ‘K26%’ OR icd_code LIKE ‘K27%’ OR icd_code LIKE ‘531%’ OR icd_code LIKE ‘532%’ OR icd_code LIKE ‘533%’ OR icd_code LIKE ‘534%’ THEN 1 ELSE 0 END) AS pud,

MAX(CASE WHEN icd_code LIKE ‘K70%’ OR icd_code LIKE ‘K73%’ OR icd_code LIKE ‘K74%’ OR icd_code LIKE ‘571%’ THEN 1 ELSE 0 END) AS liver_mild,

MAX(CASE WHEN icd_code LIKE ‘E10%’ OR icd_code LIKE ‘E11%’ OR icd_code LIKE ‘250%’ THEN 1 ELSE 0 END) AS dm,

MAX(CASE WHEN icd_code LIKE ‘E102%’ OR icd_code LIKE ‘E103%’ OR icd_code LIKE ‘E104%’ OR icd_code LIKE ‘E105%’ OR icd_code LIKE ‘E112%’ OR icd_code LIKE ‘E113%’ OR icd_code LIKE ‘E114%’ OR icd_code LIKE ‘E115%’ THEN 2 ELSE 0 END) AS dm_comp,

MAX(CASE WHEN icd_code LIKE ‘G81%’ OR icd_code LIKE ‘G82%’ OR icd_code LIKE ‘342%’ OR icd_code LIKE ‘344%’ THEN 2 ELSE 0 END) AS hemiplegia,

MAX(CASE WHEN icd_code LIKE ‘N18%’ OR icd_code LIKE ‘N19%’ OR icd_code LIKE ‘585%’ OR icd_code LIKE ‘586%’ THEN 2 ELSE 0 END) AS renal,

MAX(CASE WHEN icd_code LIKE ‘C%’ OR icd_code BETWEEN ‘140’ AND ‘172’ OR icd_code BETWEEN ‘174’ AND ‘195’ THEN 2 ELSE 0 END) AS cancer,

MAX(CASE WHEN icd_code LIKE ‘K72%’ OR icd_code LIKE ‘I85%’ OR icd_code LIKE ‘456%’ OR icd_code LIKE ‘572%’ THEN 3 ELSE 0 END) AS liver_severe,

MAX(CASE WHEN icd_code LIKE ‘C77%’ OR icd_code LIKE ‘C78%’ OR icd_code LIKE ‘C79%’ OR icd_code LIKE ‘C80%’ OR icd_code BETWEEN ‘196’ AND ‘199’ THEN 6 ELSE 0 END) AS metastatic,

MAX(CASE WHEN icd_code LIKE ‘B20%’ OR icd_code LIKE ‘B21%’ OR icd_code LIKE ‘B22%’ OR icd_code LIKE ‘042%’ THEN 6 ELSE 0 END) AS aids

FROM physionet-data.mimiciv_3_1_hosp.diagnoses_icd

GROUP BY hadm_id

),

charlson_score AS (

SELECT

hadm_id,

mi + chf + pvd + cvd + dementia + copd + rheum + pud +

CASE WHEN liver_severe > 0 THEN liver_severe ELSE liver_mild END +

GREATEST(dm, dm_comp) + hemiplegia + renal +

GREATEST(cancer, metastatic) + aids AS charlson_index

FROM charlson

),

-- SOFA scores - get first value per stay

sofa_scores AS (

SELECT stay_id, sofa_24hours AS sofa

FROM physionet-data.mimiciv_3_1_derived.sofa

WHERE hr >= 0 AND hr < 24

QUALIFY ROW_NUMBER() OVER (PARTITION BY stay_id ORDER BY hr) = 1

),

-- Orientation assessments from chartevents

orientation_events AS (

SELECT

ce.stay_id,

ce.charttime,

ce.value,

TIMESTAMP_DIFF(ce.charttime, bc.icu_intime, HOUR) AS hours_since_admission

FROM physionet-data.mimiciv_3_1_icu.chartevents ce

INNER JOIN base_cohort bc ON ce.stay_id = bc.stay_id

WHERE ce.itemid IN (229381, 223898, 228394, 228395, 228396)

AND ce.charttime BETWEEN bc.icu_intime AND TIMESTAMP_ADD(bc.icu_intime, INTERVAL 24 HOUR)

),

-- Classify orientation status

orientation_status AS (

SELECT

bc.stay_id,

CASE

WHEN EXISTS (

SELECT 1 FROM orientation_events oe

WHERE oe.stay_id = bc.stay_id

AND oe.hours_since_admission <= 6

AND LOWER(oe.value) NOT LIKE ‘%unable%’

) THEN ‘Assessed Early’

WHEN EXISTS (

SELECT 1 FROM orientation_events oe

WHERE oe.stay_id = bc.stay_id

AND oe.hours_since_admission > 6

AND oe.hours_since_admission <= 24

AND LOWER(oe.value) NOT LIKE ‘%unable%’

) THEN ‘Assessed Late’

WHEN EXISTS (

SELECT 1 FROM orientation_events oe

WHERE oe.stay_id = bc.stay_id

AND LOWER(oe.value) LIKE ‘%unable%’

) THEN ‘Unable to Assess’

ELSE ‘Never Documented’

END AS orientation_category

FROM base_cohort bc

)

-- FINAL SELECT - produces the 46,004 cohort

SELECT

bc.stay_id,

bc.age,

CASE WHEN bc.gender = ‘M’ THEN 1 ELSE 0 END AS male,

bc.hospital_expire_flag AS died,

COALESCE(sf.sofa, 0) AS sofa,

COALESCE(cs.charlson_index, 0) AS charlson,

CASE WHEN bc.admission_shift = ‘Night’ THEN 1 ELSE 0 END AS night_shift,

CASE WHEN bc.weekend_flag = ‘Weekend’ THEN 1 ELSE 0 END AS weekend,

os.orientation_category,

CASE WHEN os.orientation_category = ‘Never Documented’ THEN 1 ELSE 0 END AS never_documented,

bc.first_careunit

FROM base_cohort bc

LEFT JOIN sofa_scores sf ON bc.stay_id = sf.stay_id

LEFT JOIN charlson_score cs ON bc.hadm_id = cs.hadm_id

LEFT JOIN orientation_status os ON bc.stay_id = os.stay_id

WHERE COALESCE(sf.sofa, 0) <= 2

AND bc.first_careunit NOT LIKE ‘%Neuro%’

## REFERENCES

1. Born G. Negative telemetry in the intensive care unit: orientation assessment omission as a predictor of hospital mortality. Manuscript submitted for publication. 2026.

2. Aiken LH, Clarke SP, Sloane DM, Sochalski J, Silber JH. Hospital nurse staffing and patient mortality, nurse burnout, and job dissatisfaction. JAMA. 2002;288(16):1987–1993. PMID: 12387650

3. Needleman J, Buerhaus P, Mattke S, Stewart M, Zelevinsky K. Nurse-staffing levels and the quality of care in hospitals. N Engl J Med. 2002;346(22):1715–1722. PMID: 12037152

4. Sittig DF, Singh H. A new sociotechnical model for studying health information technology in complex adaptive healthcare systems. Qual Saf Health Care. 2010;19(Suppl 3):i68–i74. PMID: 20959322

5. Croskerry P. The importance of cognitive errors in diagnosis and strategies to minimize them. Acad Med. 2003;78(8):775–780. PMID: 12915363

6. Johnson AEW, Bulgarelli L, Shen L, Gayles A, Shammout A, Horng S, Pollard TJ, Hao S, Moody B, Gow B, Lehman LH, Celi LA, Mark RG. MIMIC-IV, a freely accessible electronic health record dataset. Sci Data. 2023;10(1):1. PMID: 36596836

7. Vincent JL, Moreno R, Takala J, Willatts S, De Mendonça A, Bruining H, Reinhart CK, Suter PM, Thijs LG. The SOFA (Sepsis-related Organ Failure Assessment) score to describe organ dysfunction/failure. Intensive Care Med. 1996;22(7):707–710. PMID: 8844239

8. Charlson ME, Pompei P, Ales KL, MacKenzie CR. A new method of classifying prognostic comorbidity in longitudinal studies: development and validation. J Chronic Dis. 1987;40(5):373–383. PMID: 3558716

9. Austin PC. An introduction to propensity score methods for reducing the effects of confounding in observational studies. Multivariate Behav Res. 2011;46(3):399–424. PMID: 21818162

10. VanderWeele TJ, Ding P. Sensitivity analysis in observational research: introducing the E-value. Ann Intern Med. 2017;167(4):268–274. PMID: 28693043

11. Kalisch BJ, Landstrom GL, Hinshaw AS. Missed nursing care: a concept analysis. J Adv Nurs. 2009;65(7):1509–1517. PMID: 19456994

12. Kalisch BJ. Missed nursing care: a qualitative study. J Nurs Care Qual. 2006;21(4):306–313. PMID: 16985399

13. Ely EW, Inouye SK, Bernard GR, Gordon S, Francis J, May L, Truman B, Speroff T, Gautam S, Margolin R, Hart RP, Dittus R. Delirium in mechanically ventilated patients: validity and reliability of the confusion assessment method for the intensive care unit (CAM-ICU). JAMA. 2001;286(21):2703–2710. PMID: 11730446

14. Bergeron N, Dubois MJ, Dumont M, Dial S, Skrobik Y. Intensive Care Delirium Screening Checklist: evaluation of a new screening tool. Intensive Care Med. 2001;27(5):859–864. PMID: 11430542

15. Johnson AEW, Pollard TJ, Mark RG. MIMIC-IV. PhysioNet. Accessed 2025. https://mimic.mit.edu/

16. von Elm E, Altman DG, Egger M, Pocock SJ, Gøtzsche PC, Vandenbroucke JP; STROBE Initiative. The Strengthening the Reporting of Observational Studies in Epidemiology (STROBE) statement: guidelines for reporting observational studies. Ann Intern Med. 2007;147(8):573–577. PMID: 17938396

17. Newman-Toker DE, Schaffer AC, Yu-Moe CW, Nassery N, Saber Tehrani AS, Clemens GD, Wang Z, Zhu Y, Fanai M, Brush DE. Burden of serious harms from diagnostic error in the USA. BMJ Qual Saf. 2024;33(2):109–120. PMID: 37460118

18. Sessler CN, Gosnell MS, Grap MJ, Brophy GM, O’Neal PV, Keane KA, Tesoro EP, Elswick RK. The Richmond Agitation-Sedation Scale: validity and reliability in adult intensive care unit patients. Am J Respir Crit Care Med. 2002;166(10):1338–1344. PMID: 12421743

19. Schubert M, Glass TR, Clarke SP, Aiken LH, Schaffert-Witvliet B, Sloane DM, De Geest S. Rationing of nursing care and its relationship to patient outcomes: the Swiss extension of the International Hospital Outcomes Study. Int J Qual Health Care. 2008;20(4):227–237. PMID: 18436556

20. Teasdale G, Jennett B. Assessment of coma and impaired consciousness: a practical scale. Lancet. 1974;2(7872):81–84. PMID: 4136544

21. Krewulak KD, Stelfox HT, Parsons Leigh J, Ely EW, Fiest KM. Incidence and prevalence of delirium subtypes in an adult ICU: a systematic review and meta-analysis. Crit Care Med. 2018;46(12):2029–2035. PMID: 30234569

22. Ely EW, Shintani A, Truman B, Speroff T, Gordon SM, Harrell FE Jr, Inouye SK, Bernard GR, Dittus RS. Delirium as a predictor of mortality in mechanically ventilated patients in the intensive care unit. JAMA. 2004;291(14):1753–1762. PMID: 15082703

23. Thomason JW, Shintani A, Peterson JF, Pun BT, Jackson JC, Ely EW. Intensive care unit delirium is an independent predictor of longer hospital stay: a prospective analysis of 261 non-ventilated patients. Crit Care. 2005;9(4):R375–R381. PMID: 16137350

24. Alzoubi KH, Rabab’h OA, Al Azzam SI, Alomari MA, Alhajlah S, Alrabadi N, Almomani B, Khabour OF, Alzoubi A. Delirium incidence, predictors and outcomes in the intensive care unit: a prospective cohort study. Int J Nurs Pract. 2024;30(1):e13154. PMID: 37051669

25. Born G. Behavioral telemetry for diagnostic error prevention: a call for real-time cognitive state monitoring in clinical workflows. J Med Internet Res. Manuscript submitted for publication. 2026.

